# External Validation of DeepBleed: The first open-source 3D Deep Learning Network for the Segmentation of Intracerebral and Intraventricular Hemorrhage

**DOI:** 10.1101/2023.02.27.23286358

**Authors:** Haoyin Cao, Andrea Morotti, Federico Mazzacane, Dmitriy Desser, Frieder Schlunk, Christopher Güttler, Helge Kniep, Tobias Penzkofer, Jens Fiehler, Uta Hanning, Andrea Dell’Orco, Jawed Nawabi

## Abstract

**Objectives:** DeepBleed is the first publicly available deep neural network model for the 3D segmentation of acute intracerebral hemorrhage (ICH) and intraventricular hemorrhage (IVH) on non-enhanced CT scans (NECT). The aim of this study was to evaluate the generalizability in an independent heterogenous ICH cohort and to improve the prediction accuracy by retraining the model.

**Methods:** This retrospective study included patients from three European stroke centers diagnosed with acute spontaneous ICH and IVH on NECT between January 2017 and June 2020. Patients were divided into a training-, validation- and test cohort according to the initial study. Model performance was evaluated using metrics of dice score (DSC), sensitivity, and positive predictive values (PPV) in the original model (OM) and the retrained model (RM) for each ICH location. Students’ t-test was used to compare the DSC between the two models. A multivariate linear regression model was used to identify variables associated with the DSC. Pearson correlation coefficients (r) were calculated to evaluate the volumetric agreement with the manual reference (ground truth: GT). Intraclass correlation coefficients (ICC) were calculated to evaluate segmentation agreement with the GT compared to expert raters.

**Results:** In total, 1040 patients were included. Segmentations of the OM had a median DSC, sensitivity, and PPV of 0.84, 0.79, and 0.93, compared to 0.83, 0.80, and 0.91 in the RM, adjusted p-values > 0.05. Performance metrics for infratentorial ICH improved from a median DSC of 0.71 for brainstem and 0.48 for cerebellar ICH in the OM to 0.77 and 0.79 in the RM. ICH volume and location were significantly associated with the DSC, p-values < 0.05. Volumetric measurements showed strong agreement with the GT (r > 0.90), p-value >0.05. Agreement of the automated segmentations with the GT were excellent (ICC ≥ 0.9, p-values <0.001), however worse if compared to the human expert raters (p-values <0.0001).

**Conclusions:** DeepBleed demonstrated an overall good generalization in an independent validation cohort and location specific variances improved significantly after model retraining. Segmentations and volume measurements showed a strong agreement with the manual reference; however, the quality of segmentations was lower compared to human expert raters. This is the first publicly available external validation of the open-source DeepBleed network for spontaneous ICH introduced by Sharrock et al.

## Introduction

Spontaneous intracerebral hemorrhage (ICH) is a major cause of morbidity and mortality worldwide despite the relatively small contribution to all stroke types of up to 27%.^1-4^ The prognosis after ICH is particularly affected by the ICH volume in addition to its location, the presence of intraventricular hemorrhage (IVH), and acute hematoma expansion (HE), a potentially modifiable risk factor.^5^ Thus, early identification and volumetric estimation on neuroimaging is a crucial step to guide further patient management, inform about surgical interventions, aid discussions of prognosis and form clinic trials.^6-9^ The ABC/2 method has remained a clinically well-established formula to manually estimate the ICH volume^10^ despite the consistent reports of under-or overestimation in large and irregular bleedings. The semiautomatic measurements of ICH and IVH are equally limited as they are labor intensive and time consuming.^11^ Novel deep learning-based models have the potential to quantify ICH and IVH volumes rapidly and accurately in a fully automated approach and are therefore in high demand.^12^ The DeepBleed network presented by Sharrock et al. is the first publicly available 3D neural network for the segmentation of ICH and IVH.^13^ Despite being trained and validated on a large dataset from the MISTIE II and III trial series,^14,15^ the generalization performance in an external, independent cohort has not been described yet. This step is of particular importance in imaging-based segmentation networks as they have increasingly shown inconsistent performance results on external datasets, with some even demonstrating a substantial performance decrease.^16^ In particular, the performance of DeepBleed in the detection and segmentation of infratentorial and small ICH remains undetermined as these two subsets were excluded from the MISTIE trials.^14,15,17^ The aim of this study was to evaluate the generalizability and further improve the robustness of the proposed DeepBleed network. Therefore, we hypothesized that the DeepBleed network would accurately detect and segment ICH and IVH regardless of its location and size. To test and evaluate this, the following threefold steps were performed: First, we validated the original DeepBleed model (OM) in an independent multicenter cohort, which included, among others, infratentorial and small ICH. Secondly, we retrained the model (RM) to test the effect on the validation accuracy. Third, we compared the interrater reliabilities between the OM and RM network and independent human raters.

## Materials and Methods

### Study population

This retrospective study was approved by the local ethics committee (Charité Berlin, Germany [protocol number EA1/035/20], University Medical-Center Hamburg, Germany [protocol number WF-054/19], and IRCCS Mondino Foundation, Pavia, Italy [protocol number 20190099462]). Written informed consent was waived by the institutional review boards. All study protocols and procedures were conducted in accordance with the Declaration of Helsinki. The study included patients of ≥ 18 years and older who were diagnosed with primary spontaneous ICH on noncontrast Computed Tomography (NECT) between January 2017 and June 2020. Patients with multiple ICH, artifacts, external ventricular drain (EVD) or any other type of surgical procedure, and secondary hemorrhage following head trauma, ischemic infarction, neoplastic mass lesions, ruptured cerebral aneurysms, or vascular malformations were excluded from the study as presented in Figure 1. Clinical data obtained from medical record included age, sex, National Institutes of Health Stroke Scale (NIHSS) and Glasgow Come Scale (GCS) at admission, symptom onset time and imaging time.

**Figure 1:**
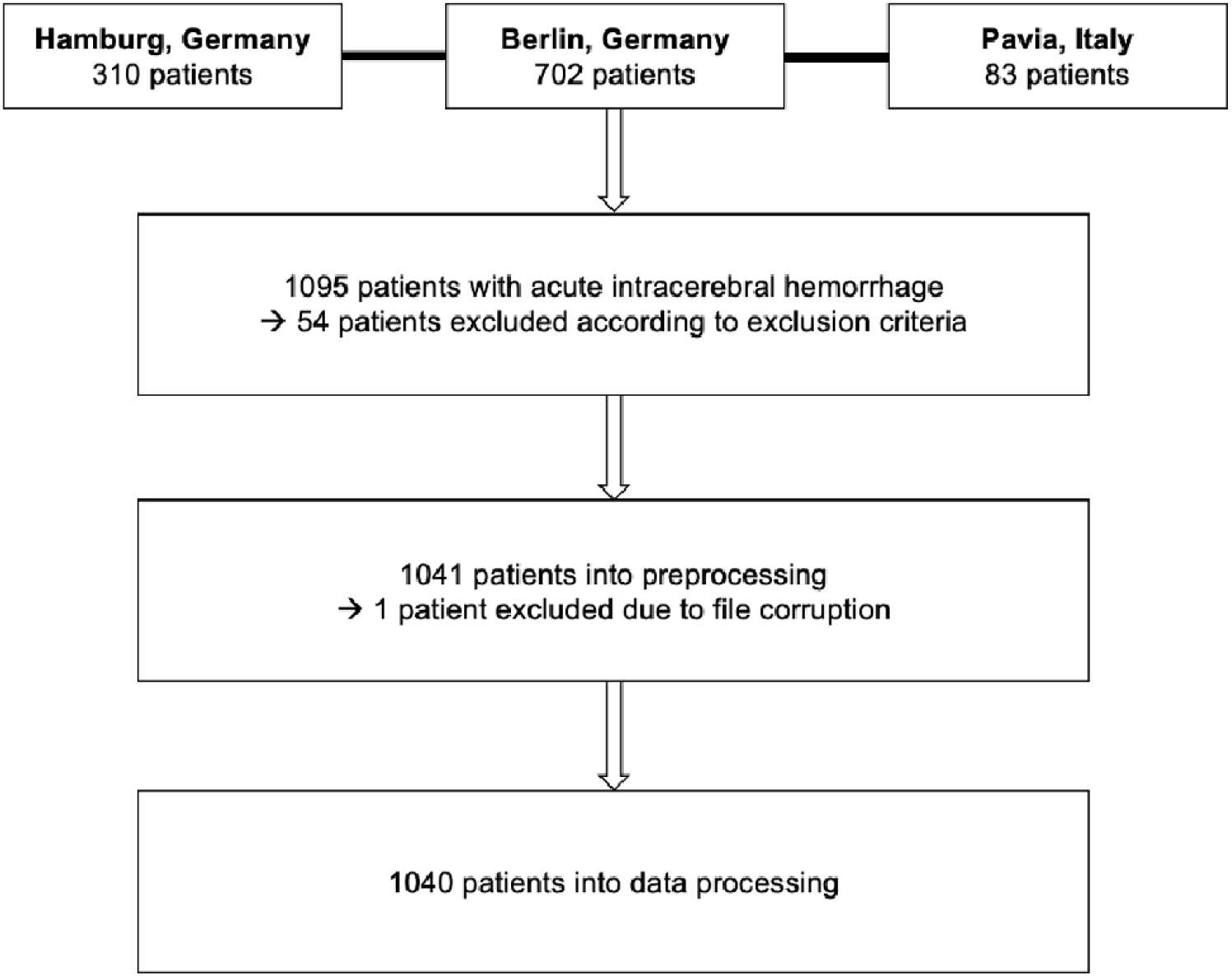
Patient flow diagram. **Legend:** Flow diagram of patients included from three European participating sites and final patient cohort after further exclusion.

**Figure 2:**
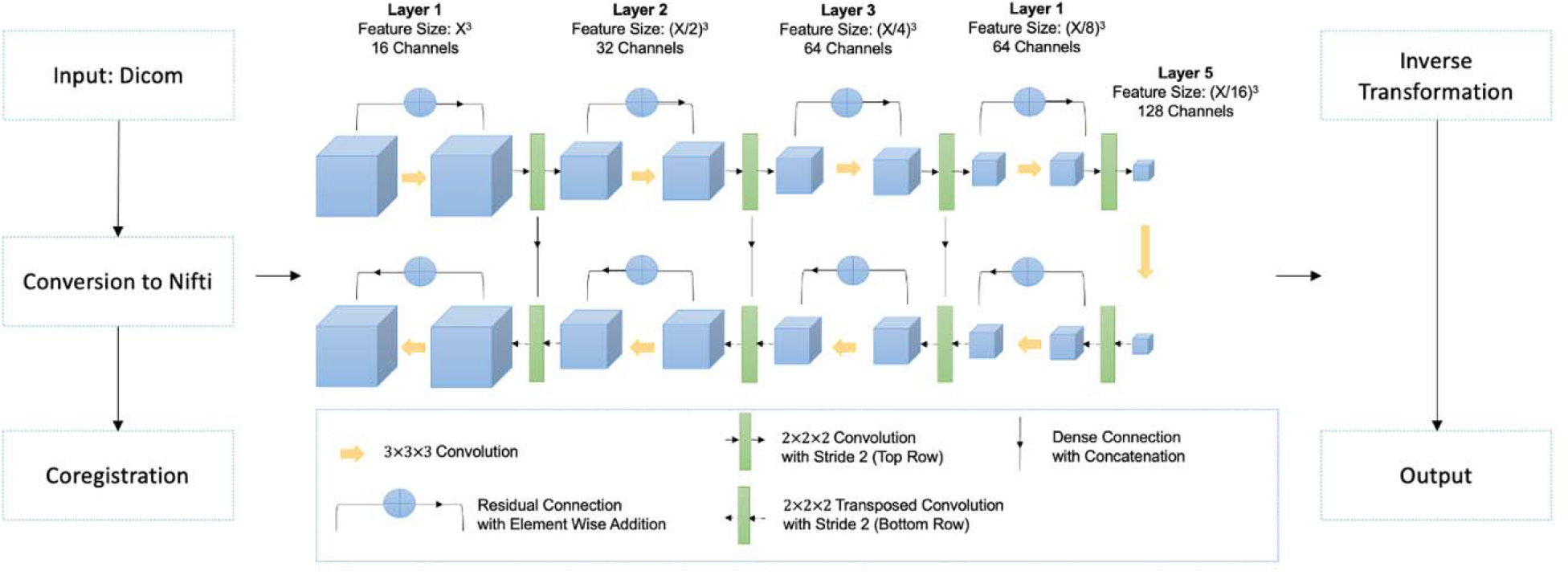
Processing pipeline of the 3D DeepBleed network. **Legend:** After gantry tilt and unequal slices were corrected, DICOM data was converted using NIfTI. For brain extraction and coregistration, Python preprocessing pipelines were used. DeepBleed was then used to predict intracerebral hemorrhage (ICH) and intraventricular hemorrhage (IVH). In the final step, the predictions from the previous template registration were inversely transformed into the native space.

### Image acquisition and manual segmentation

Participating sites acquired NECT images according to their local imaging protocols. Anonymized imaging data were retrieved from the local picture archiving and communication system (PACS) servers and converted to Digital Imaging and Communications in Medicine (DICOM) format according to local guidelines. DICOM data were then transformed to Neuroimaging Informatics Technology Initiative (NifTI) for further imaging analysis. Images were analyzed for the presence of IVH and the ICH location by one experienced neuroradiologist (J.N., 5 years of experience in ICH imaging research). Supratentorial bleedings in cortical and subcortical location were classified as lobar and hemorrhages involving the thalamus, basal ganglia, internal capsule and deep periventricular white matter as deep.^18^ Infratentorial bleedings were classified within the brainstem and pons or cerebellum.^19^ Ground truth (GT) masks of ICH and IVH were manually segmented on CT scans by two experienced raters (both 3 and 5 years of experience in ICH imaging research) who were supervised by one neuroradiologist (J.N.) who inspected each ICH and IVH mask for the quality of segmentations, and corrected them if necessary. Segmentation of the ICH and IVH was performed using the ITK-SNAP software version 3.8.0.^20^ Because DeepBleed does not differentiate ICH and IVH, fusion masks comprising ICH and IVH were created for all subjects with IVH presence. This step was followed by a data quality control of the generated fusion masks by a fourth rater (H.C., 3 years of experience in ICH imaging research) and corrected by one computer scientist, if necessary (A. D’O., 5 years of experience in neuroimaging informatics). A repeated segmentation was performed 3 months apart by two expert raters for 60 patients, selected in the test set, to calculate inter-reader agreement as described below (J.N. and F.M., 5 years of experience in ICH imaging research). All readers independently analyzed and segmented images in a random order, blind to all demographic data and were not involved in the clinical care of assessment of the enrolled patients.

### Pre- and postprocessing

The preprocessing comprised two steps as described in the original study: The brain extraction and the coregistration. We used the same code released from the original authors. Briefly, after setting to zero the CT scan intensities lower than 0 or higher than 100, the brain extraction was performed with FSL Brain Extraction Tool (BET)^21^ setting the fractional intensity parameter (−f flag) to 0.01. The rigid coregistration was performed with ANTs^22^ using a 1.5 mm³ isotropic CT template^23^. The resulting transformation was applied to the GT masks as well. As postprocessing, a threshold of 0.6 was applied to the resulting probability maps, setting the higher values to 1 and the others to 0. Finally, the inverse coregistration of the resulting mask was performed using the inverse transformation matrix.

### Model Retraining

In addition to the general exclusion criteria described in the above, the following additional exclusion criteria were applied, to the training cohort as they might negatively influence the model: Symptom onset > 24 hours prior to the admission CT or an unknown time of symptom onset as well as an admission ICH volume > 30 mL. As described in the original study, Adaptive Moment Estimation (Adam) was used as the optimization function,^24^ dice similarity coefficient (DSC) was used as a loss function,^25^ and 100, randomly selected subjects were used. After testing various combinations, the optimal learning rate was 1×10^−4^ and a batch size 3 was chosen. Initially, the train dataset was shuffled; we stopped the training if, after the first 10th epoch, the current epoch did not improve, and validation was performed every five epochs.

### Model Testing

For the model testing, pre- and postprocessing described as in the above were applied to the OM and RM test dataset. Contrary to the training dataset, test dataset was defined according to the original criteria of our study with no additional exclusion criteria applied.

### Code availability

The code is publicly available as Jupiter Notebooks on GitHub with the following link: https://github.com/Orangepepermint/retraindeepbleed. It is written in Python v3.9^26^ using the following libraries: NumPy v1.23.3,^27^ FSLpy v3.9.0,^28^ and ANTsPy v0.3.1.^22^

### Statistical Analysis

Statistics were conducted using GraphPad Prism v9.0.2^29^ and R (the R project for statistical computing) using tidyverse.^30^ Various metrics have been used to evaluate the DeepBleed performance and compare the segmentations from our RM with the OM. These include DSC, sensitivity, positive predictive value (PPV), and volume measurements. T-tests were used to compare the DSC, sensitivity, and PPV distributions between the OM and RM. To determine factors to influence the segmentation performance a linear regression model with the following formula was conducted:

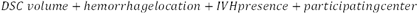

Pairwise correlations among volumes measured from each of the three segmentation methods (GT masks, OM and RM DeepBleed network) were assessed using the Pearson correlation coefficient (r). Agreement between two raters and the OM and RM DeepBleed network were assessed using the intraclass correlation coefficient (ICC) on the DSC. Moreover, a repeated measures ANOVA was performed with a pairwise t-test as a post-hoc. Cohen’s d effect size was determined as-well. A p-value<0.05 was considered significant. Bonferroni adjustment was applied where necessary. Adjusted p-values are indicated as p_adj_-value.

## Results

### Demographics and Characteristics of the Study Cohort

The manual review of the images led to an elimination of 54 patients due to exclusion criteria and manual segmentation errors. The final dataset was composed of 1040 patients, NECT scans and respective masks. The mean age was 69.6 (SD 14.2) years. The median NIHSS and GCS score were 7.5 (IQR 1-13) and 13 (IQR 8-15), respectively. Imaging was performed within a median symptom onset time of 4.3 hours (IQR 1.5-15.1). 519 patients (49.9%) presented with an IVH with a mean combined ICH and IVH volume of 74.9 (SD 41.6) ml. 521 patients (50.1%) presented with ICH only with a mean volume of 41.5 (SD 32.8) ml. The final training to validation to test ratio was 5:1:46. No significant differences of demographic characteristics were found between training, validation, and test subjects as presented in Table 1.

**Table 1:** Comparison of the demographics and volumetry. Demographics and descriptive characteristics have been compared between the training, validation, and test datasets. NIHSS, National Institutes of Health Stroke Score; GCS, Glasgow Coma Scala; ICH, intracerebral hemorrhage; IQR, interquartile range; IVH, intraventricular hemorrhage; SD, standard deviation;. ^1^ one-way-ANOVA test.

| Variable | Training cohort<br>(n=100) | Validation cohort<br>(n=20) | Test cohort<br>(n=920) | p-value<sup>1</sup> |
| --- | --- | --- | --- | --- |
| Age [years], mean ± SD | 70.5 ± 13.1 | 68 ± 13.4 | 69.6 ± 14.2 | 0.84 |
| Gender, n (%) |  |  |  |  |
| Male | 41 (41) | 11 (55) | 516 (56) |  |
| Female | 59 (59) | 9 (45) | 404 (44) |  |
| NIHSS score, median<br>(IQR) | 7.5 (4-14) | 10 (5-15) | 7 (8-13) | 0.70 |
| GCS score, median (IQR) | 13 (8-15) | 14 (11-15) | 13 (7-15) | 0.16 |
| Symptom onset to imaging | 4.3 (1.5-15.1) | 3.9 (1.8-9.4) | 4.23 (1.8-14.6) | 0.99 |
| **[hours], median (IQR)** |  |  |  |  |
| **ICH location, n (%)** |  |  |  | 0.17 |
| Lobar | 44 (44) | 6 (30) | 338 (36.7) |  |
| Deep | 46 (46) | 7 (35) | 455 (49.5) |  |
| Brainstem | 3 (3) | 3 (15) | 41 (4.5) |  |
| Cerebellum | 7 (7) | 4 (20) | 86 (9.3) |  |
| **ICH + IVH volume [ml],** |  |  |  |  |
| **mean ± SD** | 83.8 ± 47.2 | 56.4 ± 89.7 | 76.5 ± 44.9 | 0.97 |
| **ICH volume [ml], mean ±** |  |  |  |  |
| **SD** | 27.7 ± 30.2 | 34.2 ± 30.9 | 44.1 ± 14.2 | 0.43 |
| **IVH volume [ml], mean ±** |  |  |  |  |
| **SD** | 61.1 ± 49.1 | 22.2 ± 77.1 | 34.5 ± 42.5 | 0.87 |

### Model Retraining and Testing

With a Nvidia RTX 3090 GPU, the model was trained for 810 epochs in 16 hours. Illustrative examples of segmentations are displayed in Figure 3. Model performance metrics of the OM and RM derived in the test set are presented in detail in Table 2 with additionally DSC metrics illustrated in Figure 4A. The overall DSC for the segmentations across all locations were relatively similar in both DeepBleed models with a median DSC of 0.84 (95% CI 0.73-0.88) in the OM and 0.83 (95% CI 0.74-0.88) in the RM. DSC given separately for each location were also almost equally high in supratentorial ICH with a median DSC of 0.86 (95% CI 0.80-0.89) in deep ICH and 0.84 (95% CI 0.78-0.89) in lobar ICH in the OM compared to 0.87 (95% CI 0.81-0.90) and 0.83 (95% CI 0.72-0.88) in the RM, respectively. In comparison, performance metrics in infratentorial locations demonstrated an overall lower DSC with a median DSC of 0.71 (95% CI 0.46-0.78) in cerebellar and 0.48 (95% CI 0.23-0.64) in brainstem ICH in the OM. DSC metrics improved in the RM, especially in cerebellar ICH, with a median DSC of 0.79 (95% CI 0.65-0.84) and 0.77 (95% CI 0.57-0.83) in brainstem ICH. OM and PM performance metrics were significantly different for the DSC and sensitivity, p_adj_-values < 0.0001, compared to the PPV, p_adj_-value > 0.05.

**Table 2:** Original and retrained model performance metrics. Model Performance across the original (OM) and retrained (RM) DeepBleed models. All datasets and hemorrhage locations were evaluated for dice scores (DSC), sensitivity, and positive predictive value (PPV) and are given as median with 95% confidence intervals. The DSC of the original (OM) and retrained weights (RM) were compared using t-tests with adjusted p-values. 95%CI, 95% confidence interval; padj-value; adjusted p-value. ^1^ = paired t-test between OM and RM for the specified metric.

| Metric | All locations | Deep | Lobar | Brainstem | Cerebellar |
| --- | --- | --- | --- | --- | --- |
| **OM** |  |  |  |  |  |
| DSC | 0.84 (0.73, 0.88) | 0.86 (0.80, 0.89) | 0.84 (0.78, 0.89) | 0.71 (0.46, 0.78) | 0.48 (0.23, 0.64) |
| Sensitivity | 0.79 (0.65, 0.86) | 0.85 (0.79, 0.91) | 0.80, (0.70, 0.87) | 0.58 (0.38, 0.74) | 0.34 (0.13, 0.49) |
| PPV | 0.93 (0.85, 0.97) | 0.91 (0.85, 0.95) | 0.99 (0.85, 0.97) | 0.88 (0.76, 0.94) | 0.94 (0.76, 0.99) |
| **RM** |  |  |  |  |  |
| Dice Score | 0.83 (0.74, 0.88) | 0.87 (0.81, 0.90) | 0.83 (0.72, 0.88) | 0.77 (0.57, 0.83) | 0.79 (0.65, 0.84) |
| Sensitivity | 0.80 (0.69, 0.87) | 0.85 (0.79, 0.91) | 0.79 (0.63, 0.88) | 0.72 (0.57, 0.79) | 0.75 (0.59, 0.84) |
| PPV | 0.91 (0.84, 0.95) | 0.91 (0.85, 0.95) | 0.92 (0.63, 0.88) | 0.87 (0.77, 0.94) | 0.88 (0.79, 0.94) |
| **Comparison** |  |  |  |  |  |
| **OM and RM** | **t<sup>1</sup>** | **p<sub>adj</sub>-value** |  |  |  |
| DSC | -5.9 | 0.001 |  |  |  |
| Sensitivity | 1.45 | > 0.05 |  |  |  |
| PPV | -7.23 | 0.001 |  |  |  |

**Figure 3:**
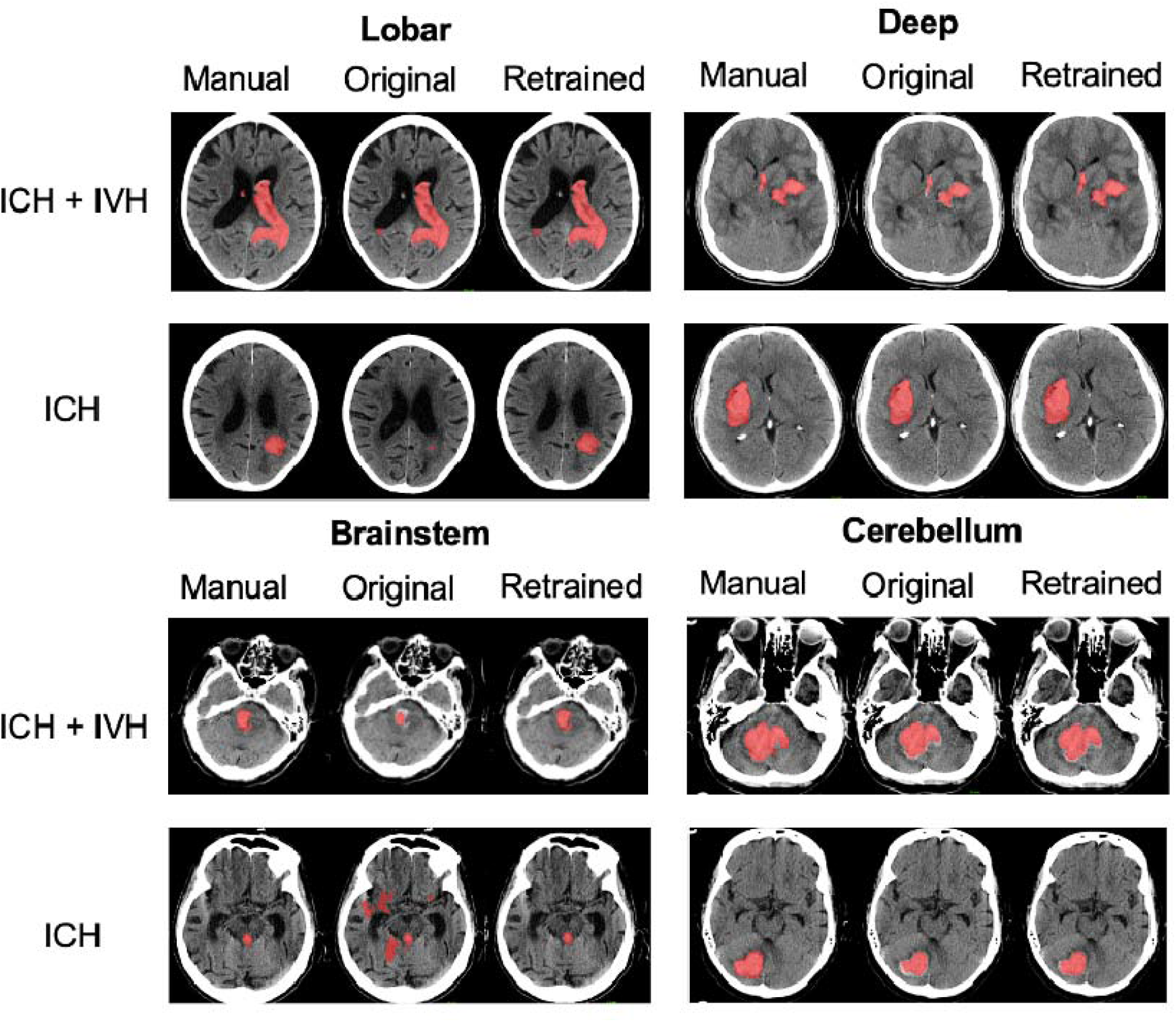
Segmentations of the original and retrained model across different locations. **Legend:** Illustrative examples of ground truth segmentations (red) for intracerebral hemorrhage (ICH) with intraventricular hemorrhage (IVH; upper rows) and ICH only (lower rows) given for deep, lobar, brainstem, and cerebellar ICH. Segmentations given are displayed for manual segmentations (left column), and DeepBleed segmentations with the original model (OM, middle column), and the retrained model (RM, right column).

**Figure 4:**
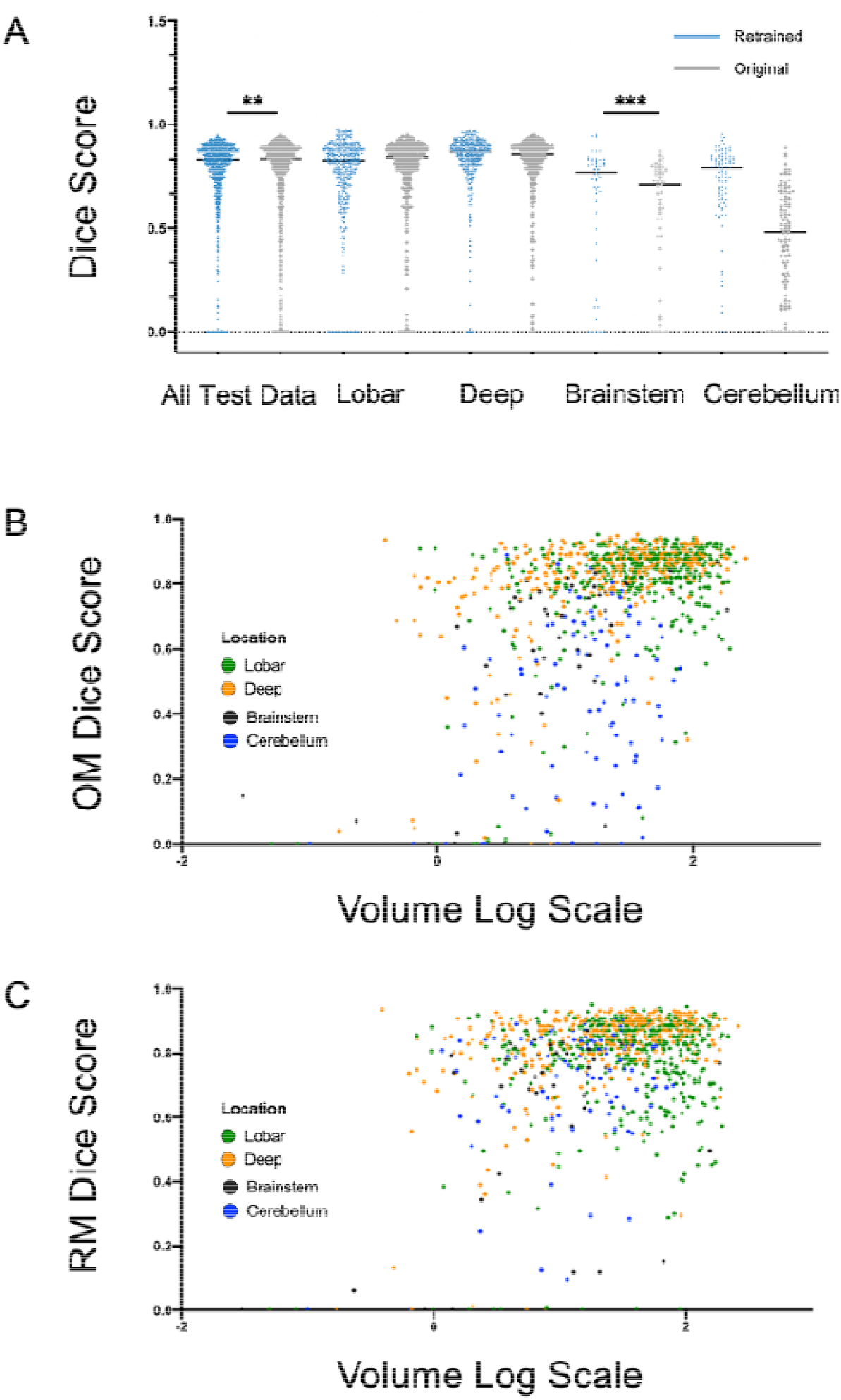
Model performance of the original and retrained model across different locations. **Legend:** A: Comparison of dice scores across lobar, deep, brainstem and cerebellar hemorrhage with original weights (OM, grey) and retrained weights (RM, blue). B-C: Relationship between hemorrhage volume and dice scores across different locations. B: Automatic segmentation with OM. C: Automatic segmentation with RM.

### Analysis of factors influencing the model performance

The results from the linear models are summarized in Table 3. Overall model performance was negatively influenced by the ICH location. However, deep ICH was the only location that was not significantly associated with a DSC loss performance neither in the OM or RM network, p-values > 0.05. While the slope coefficients for lobar ICH remained relatively similar in the OM and RM (−0.04, SE 0.02 and -0.05, SE 0.02), the negative effect of brainstem and cerebellar ICH decreased from -0.19 (SE 0.03) and -0.13 (SE 0.02) in the OM to -0.18 (SE 0.03) and -0.09 (SD 0.02) in the RM, p-values < 0.001. ICH volume increase had a strong positive effect on the DSC in the RM network, whereas the inverse effect on the DSC was found in the OM. The presence of IVH and the data originating site had no significant effect on the DSC in the OM or RM. The correlation of ICH location and volume on the DSC are illustrated in Figure 4 B-C.

**Table 3:**
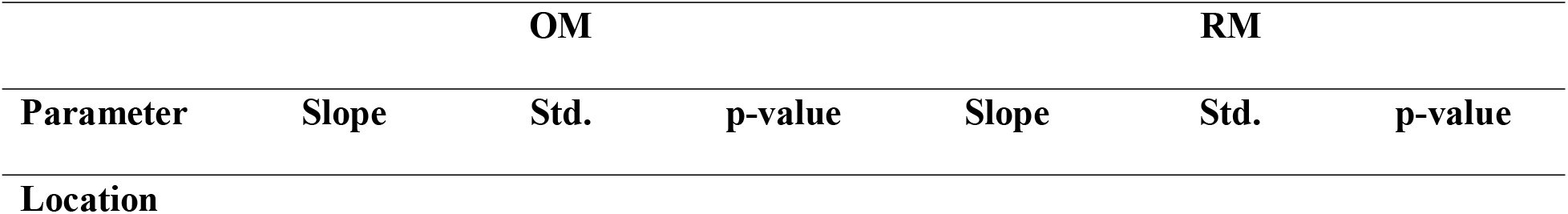
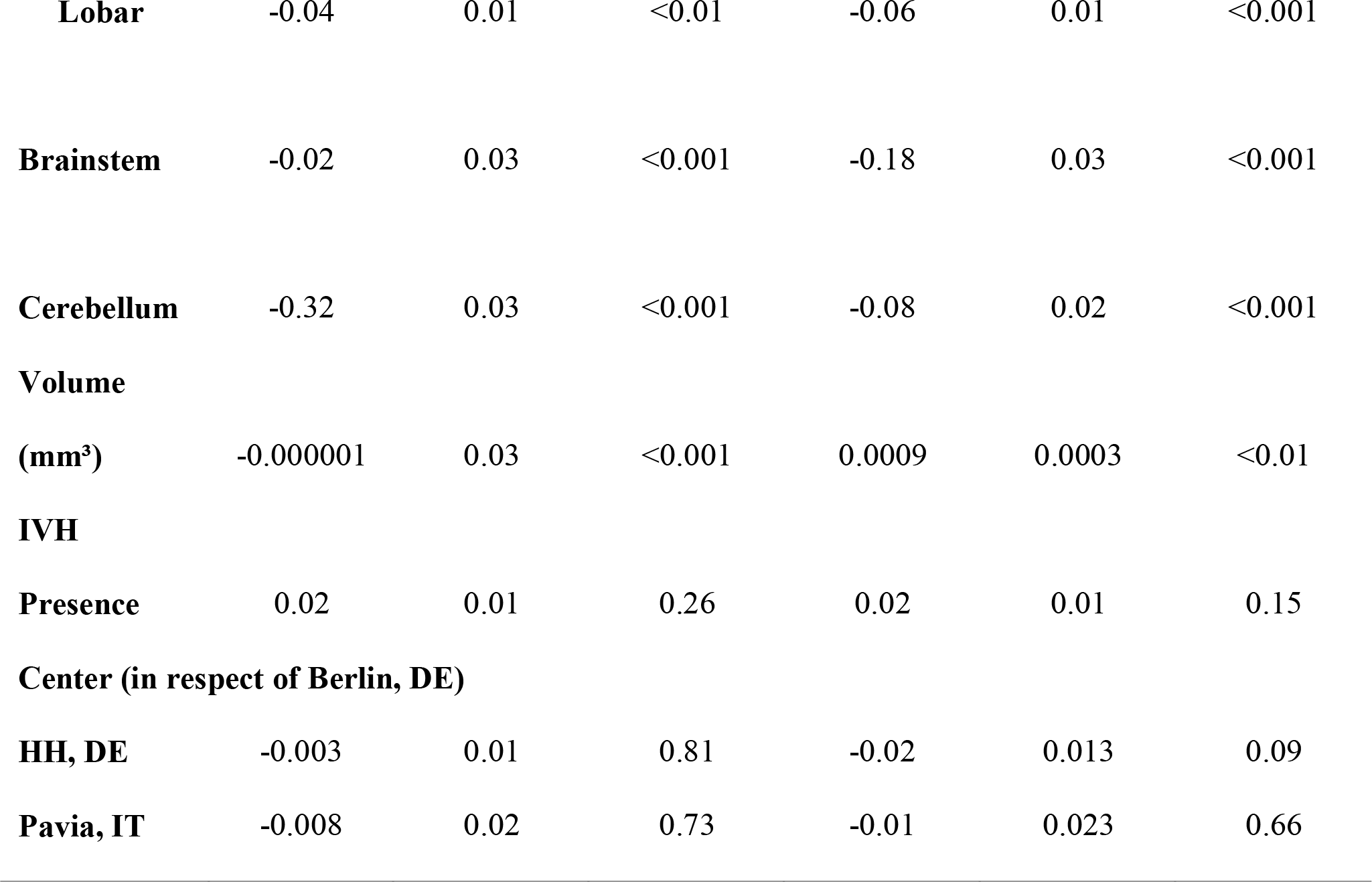
Factors influencing the original and retrained model performance. Linear regression analysis on variables influencing the model performance in the original (OM) and retrained (RM) Deepbleed model. DE, Germany; HH, Hamburg; IT, Italy; IVH, intraventricular hemorrhage; STD, standard deviation.

### Volume and segmentation agreement analysis

Figure 5A shows the correlation between the GT masks and DeepBleed’s automatic volume prediction with the OM and RM. Overall strong correlations were observed among the three segmentation methods (r > 0.9, p-value < 0.001), however, correlations among both DeepBleed models, OM and RM, were highest, whereas their correlation with GT volumes was lowest (r = 0.92 for OM and r = 0.94 for RM). The median volumes of the three methods are displayed in Figure 5B. Repeated-measure ANOVA showed no significant effect between volume estimation of GT masks and automatic volume estimations in the DeepBleed OM and RM (F = 0.45, p-value > 0.05).

**Figure 5:**
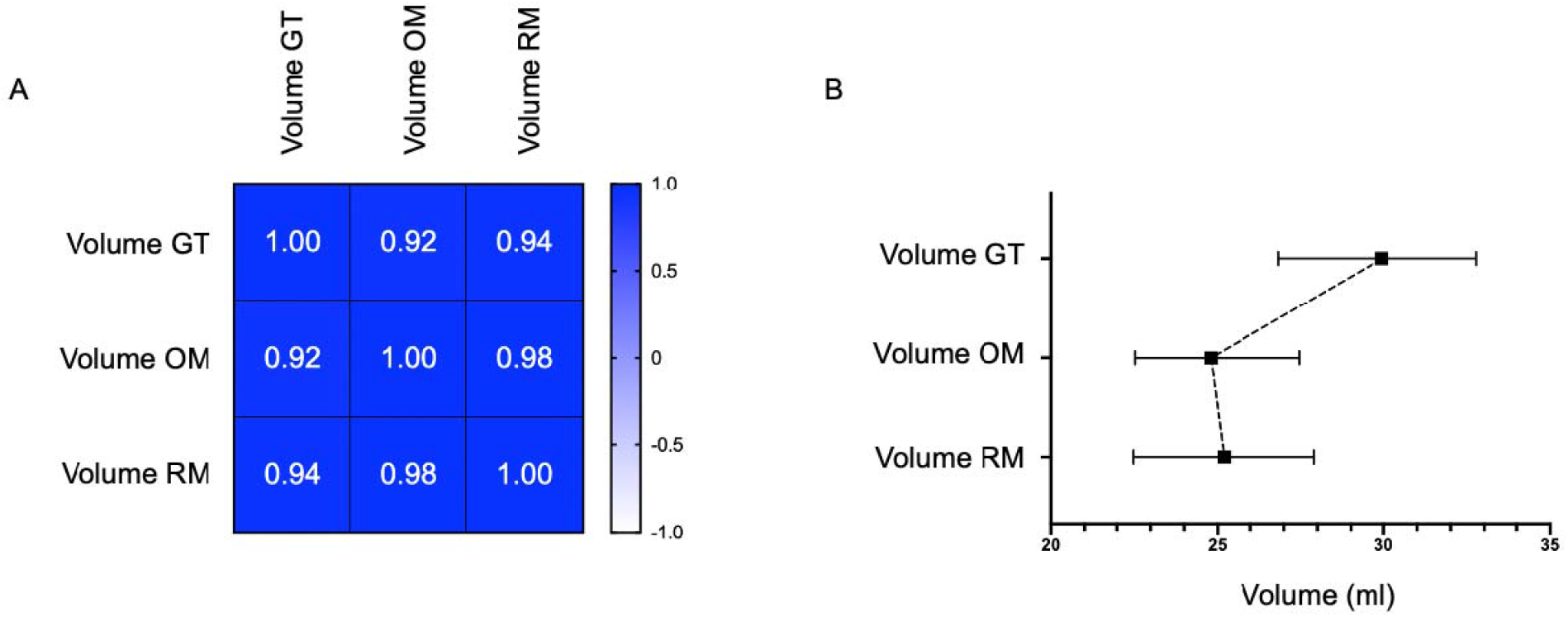
Volume agreement analysis of the original and retrained model with the ground truth. **Legend:** A: Pearson’s correlation matrix of agreement for ground truth volume and automatic segmentations of DeepBleed with original weights (original model, OM) and retrained weights (retrained model, RM). B: Median intracerebral hemorrhage and intraventricular hemorrhage volumes of ground truth and automatic segmentation with OM and RM given with 95% confidence intervall It is possible to see that deebbleed normally underestimate the Volume, probably due the probability-map threshold.

Significant agreements were found between the DSC of manual segmentations by two expert raters and the OM and RM DeepBleed network with the GT masks (ICC = 0.90 and ICC = 0.94, p-value < 0.0001) and are presented in Figure 6A-B. Repeated-measure ANOVA showed a significant effect of the rater and OM and RM on the DSC (F = 14.38, p < 0.0001). Post-hoc test showed a significant effect between OM and RM (t = 2.78, p_adj_-value < 0.05, d = 0.4, small), OM and both raters (t = -5.11 and t = –5.37, p_adj_-value <0.001 and d = 0.7, moderate for both) and RM and both rater (t = -4.9 and t = -5.3, p_adj_-value < 0.001 and d = 0.7, moderate for both). No significant difference was found between the two raters (t = -1.57, p-value > 0.05, d = 0.2, small).

**Figure 6:**
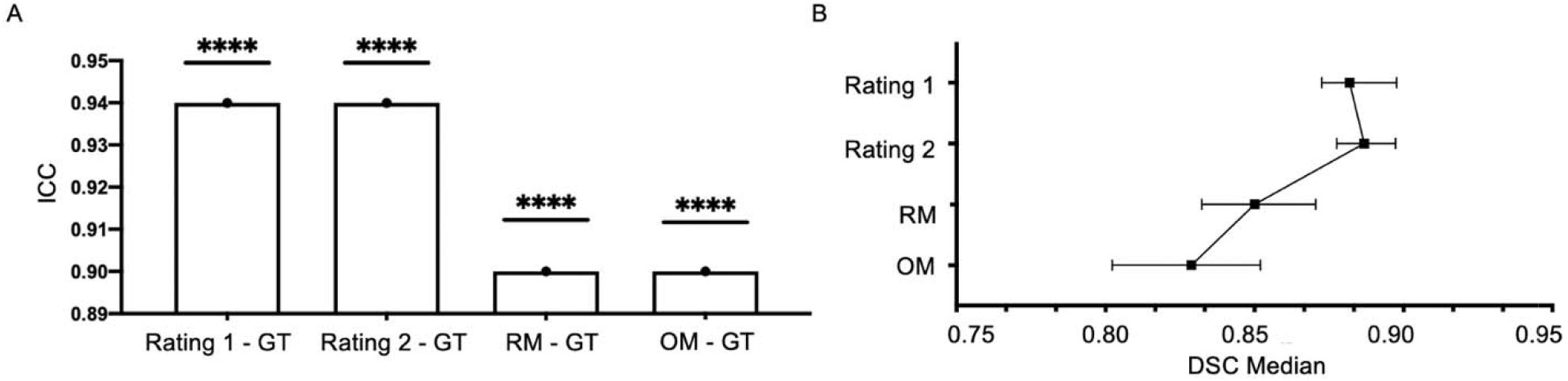
Segmentation agreement analysis of the original and retrained model and human expert raters. **Legend:** A: Intraclass Correlation Coefficient (ICC) of Dice Score (DSC) from two experienced raters, automatic segmentations of DeepBleed with original weights (OM) and retrained weights (RM) compared to ground truth (GT) of a senior stroke imaging experienced neuroradiologist. B. Median with 95% CI of DSC from each group. DSC, dice score; ICC, Intraclass Correlation Coefficient; OM, original model; RM, retrained model. **** = p-value < 0.0001.

## Discussion

Our study aimed to validate and improve the first open-source 3D deep-learning network to automatically detect and segment spontaneous ICH with IVH on CT. The overall performance of the proposed OM showed good results in our external multicenter cohort. However, the OM demonstrated significant performance differences in comparison to the RM. After our retraining, improved performance metrics were observed in infratentorial ICH, particularly in cerebellar ICH with a 1.7-fold increase of the DSC. The importance of this result was emphasized its negative effects on the DSC in our linear model which improved with a slope increase of 25% for cerebellar ICH after retraining. In comparison, model performance in supratentorial ICH was overall good in both the OM and RM. Especially deep ICH demonstrated the best and stable performance metrics and was the only location to have no negative effect on the DSC. Correspondingly, lobar ICH, as the supratentorial counterpart of deep ICH, demonstrated the second-best performance metrics and was equally stable before and after model retraining.

Counterintuitively to the latter findings, lobar ICH was negatively associated with the DSC performance. This discrepancy may be attributed to the findings on neuroimaging in which lobar ICH generally tend to present with highly irregular margins and internal density heterogeneity, contrary to the often homogenously dense and well margined deep ICH.^31^ To create a segmentation mask and perform a volumetric analysis from the above mentioned ICH, DeepBleed performs a binary prediction at the voxel level according to a predetermined threshold.^13^ With the lobar ICH phenotype in mind, this probability estimation may not translate well into an accurate segmentation and volumetric estimation as it utilizes a voxel-set threshold. However, estimating a better threshold from a probability, using a softmax, could further improve the performance in bleedings with a hypodense, non-coagulated, part which in turn is a part of the whole ICH. ^13,32^ A network that successfully implemented a softmax output is the nnU-Net network architecture.^33,34^ A prime example for the segmentation of spontaneous ICH with a nnU-Net has recently been presented by Zhao et al.^35^ The findings of this study not suggest an accurate but also ICH location independent model performance. ^35^ Despite their promising results, the DeepBleed network offers two main strengths: Although great efforts have been geared towards deep learning-based tools for ICH segmentation, the DeepBleed network has remained the only publicly available deep learning-based network for ICH segmentation. Secondly, most of the previous studies were single center based, and thus require generalization testing.^35-39^ In comparison, the original DeepBleed network was multicenter curated with data from the MISTIE II and III trial series that were conducted at 78 sites in North America, Europe, Australia, and Asia with over 500 patients included.^14,15^ In line with this, DSC metrics were independent from the participating site’s dataset in our study. Similar effects were observed for the presence of IVH and therefore, confirm the model’s robustness to scanner acquisition differences and neuropathological variability, such as the presence of IVH. Regarding the volume of the ICH, we found that a volume increase had a significantly positive effect on the DSC. This finding is consistent with previous studies that showed a positive correlation of the lesion size with the DSC.^40,41^ Therefore, the limited performance of DeepBleed in small ICH seems as no isolated case, but rather a general limitation of deep learning-based networks for lesion segmentation. The DSC has especially shown to vary in small ICH (< 5mL), although the absolute volume difference may be small.^13^ Since the DSC evaluates the quality of the alignment, which denotes the overlap between the predicted segmentation and the GT segmentation, aspects of volumetric similarity could be of more clinical importance.^42^ ICH volume is an established predictor of poor functional outcome in clinical practice and moreover, used as an inclusion and primary outcome criterion in many past and ongoing ICH trials.^14,15,17,43-45^ Our post-hoc analysis demonstrated that the automatic segmentations yielded a statistically lower agreement on the DSC compared to our human expert-raters, while still being overall excellent with an obtained ICC of > 0.9.^46,47^ In comparison, the high correlation between the manual volume and automatic volume prediction showed an underestimation of 5 mL in the automated approach which did not reach a statistical difference. This finding does not only agree well with the initial study, but also other studies for automated ICH segmentation with reported average volume errors of 2 to 5 mL and therefore, all lower than in the ABC/2 method.^12,13,48^ In addition to DeepBleed’s original design purpose for ICH trial use, the improved model could also help to inform surgical interventions and aid outcome prediction^19,49^. Especially the consideration of infratentorial ICH is of clinical importance as overall prognosis is worse compared to those located supratentorially.^19,50^ Furthermore, not only the extension of ICH into the ventricular system, but more so its intraventricular expansion has increasingly been gaining interest as a potentially modifiable factor. ^51,52,53,54^

To conclude, the DeepBleed network generalized well in an external validation cohort and location specific variances improved significantly after model retraining. Volumetric analysis showed a strong agreement with manual segmentations of expert raters, while limitations of the segmentation accuracy were illustrated yielding statistical differences with the ground truth. To our knowledge, this is the first publicly available external validation of the open-source DeepBleed network. The code and RM weights are available online^55,56^. The refined segmentation and volumetric analysis infratentorial locations highlight the generalizability of the proposed model introduced by Sharrock et al. Further studies are to improve the model are planned by including a higher number of smaller and irregularly shaped hematoma as well as extracerebral hematoma.^57^

## Data Availability

All data produced in the present study are available upon reasonable request to the authors

## Acknowledgments

Jawed Nawabi is grateful for being supported by the Berlin Institute of Health (Digital Clinician Scientist Grant funded by Charité - Universitaetsmedizin Berlin, the Berlin Institute of Health and the German Research Foundation, DFG). Frieder Schlunk was supported by the Berlin Institute of Health (Clinician Scientist Grant). Tobias Penzkofer was supported by the Berlin Institute of Health (Clinician Scientist Grant, Platform Grant), Ministry of Education and Research (BMBF, 01KX2021, 68GX21001A), German Research Foundation (DFG, SFB 1340/2), Horizon 2020 (952172). Federico Mazzacane was supported by the Italian Ministry of Health (Ricerca Corrente 2022-2024).

## Supplementary material

### Effect analysis of factors influencing the Dice Score

ANCOVA analysis on the OM showed that hemorrhage volume had an effect on the DSC (F=39.8, p<0.0001), PPV (F=4.8, p<0.0001) and sensitivity (F=4.6, p<0.0001). Only the DSC was significantly influenced by location (F=39.9, p<0.0001). There was no significant effect of IVH presence on metrics, while database had a significant effect on PPV (F=6.8, p<0.001).

The same statistical test on the RM showed a significant effect of hemorrhage volume on DSC (F=12.3, p<0.001) and PPV (F=29.5, p<0.0001) but not on sensitivity. DSC (F=14.3, p<0.0001), sensitivity (F=20.0, p<0.0001), but not PPV, were significantly affected by hemorrhage location. Furthermore, IVH presence had no effect on any metric, while the participating center only affected PPV (F=19.1, p<29.5).

**Supplementary table 1:**
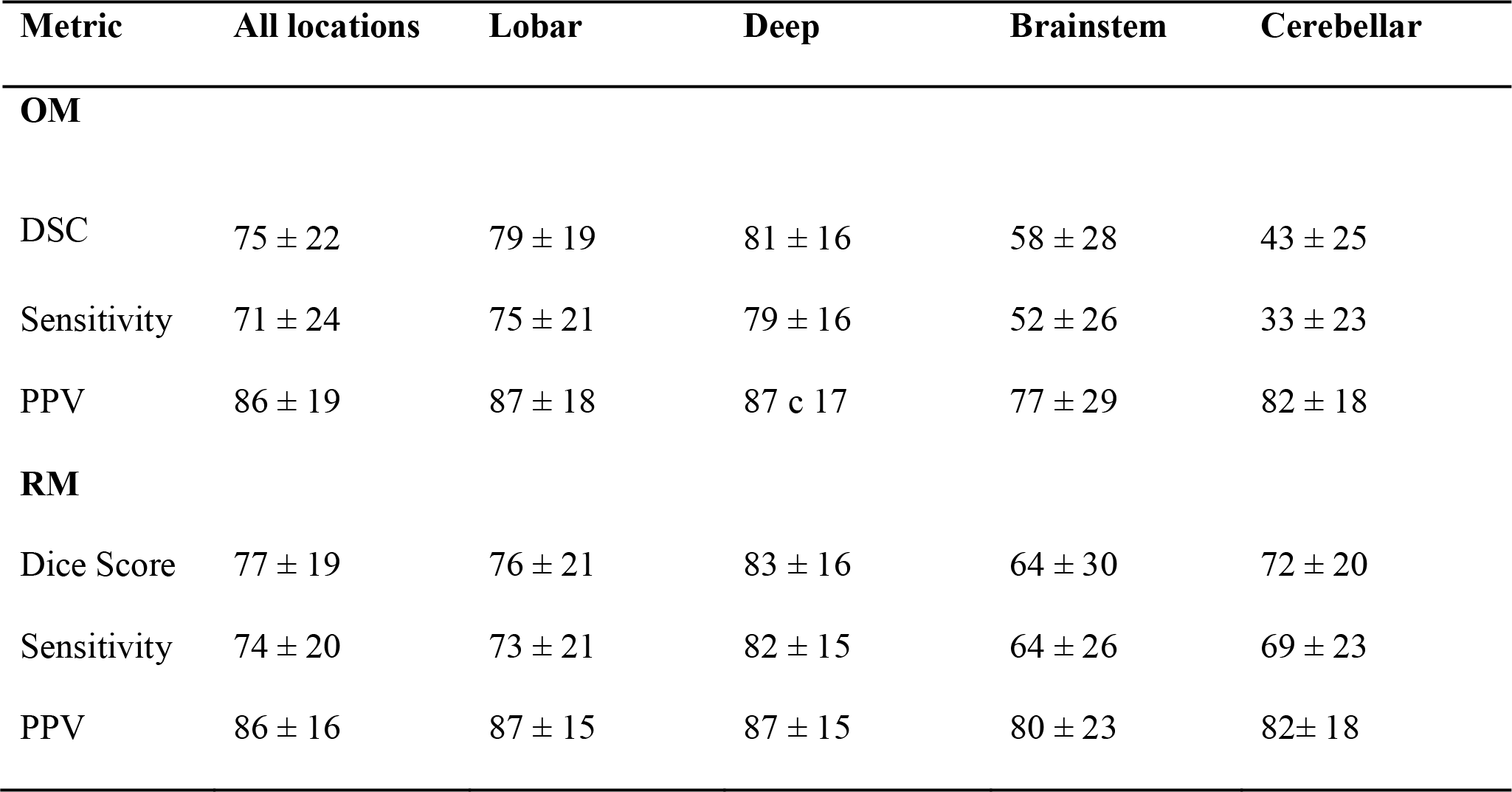
Factors influencing the original and retrained model performance. Model Performance across the original (OM) and retrained (RM) DeepBleed models. All datasets and hemorrhage locations were evaluated for dice scores (DSC), sensitivity, and positive predictive value (PPV) and are given as mean with standard deviation.

